# Validation of a blood test for multi-cancer risk stratification in a lung cancer screening cohort

**DOI:** 10.1101/2025.11.20.25340518

**Authors:** Johannes F. Fahrmann, Ehsan Irajizad, Hamid Rudsari, Jody Vykoukal, Iakovos Toumazis, Sara Khorami-Sarvestani, Sara Ansari, Jane Yang, Nicole Kettner, Jennifer B. Dennison, Edwin Ostrin, Samir Hanash

## Abstract

**Purpose:** We report a blinded validation study of a ten-protein marker blood test for assessing risk of developing or harboring nine common cancers in a prospective lung cancer screening cohort.

**Patients and Methods:** The foundation of the multi-cancer risk stratification test (MCaST), test is a 4-marker protein panel consisting of ProSFTPB, CEA, CA125, and CYFRA-21 which has been extensively validated for risk of lung cancer and which has been expanded to include six additional markers to encompass, in addition to lung cancer, prostate, colorectal, breast, ovarian, pancreatic, liver, esophageal, and stomach cancers. Blinded validation samples consisted of 1,235 plasmas collected prior to diagnosis from 171 cancer cases and 526 randomly selected non-case controls. Fixed individualized combination rules as well as cancer-specific risk thresholds predefined based on established clinical guidelines were applied.

**Results:** At the subject level, the MCaST tested positive for imminent risk of lung cancer in 65 of 73 lung cancer cases, including 45 of 51 early-stage cases, with a median time of 12.7 months (interquartile range [IQR]: 1.7 – 27.8 months) between the first positive MCaST and diagnosis of lung cancer. The positive rate of MCaST for other cancers was 88.9% for prostate, 63.6% for invasive breast, 85.7% for CRC, 50% for ovarian, 50% for pancreatic, and 67% for esophagus plus stomach cancers with a median (IQR) time of 20.7 months (10.2 – 42.8 months) from the first positive MCaST to a clinical diagnosis across the cancer types. Overall accuracy for tissue-of-origin (TOO) signal was 94.6%, and as high as 98.1% for lung cancer.

**Conclusion:** MCaST has utility for assessing cancer risk for common and lethal solid cancers in this smoker population.

## Introduction

Screening to detect cancer at an early stage is currently available in the United States for lung, breast, colon, cervical and prostate cancers.^1^ Screening is also available for gastric cancer in Asia where the incidence is high.^2^ For these cancer types, screening is generally recommended for individuals within a certain age range and in the case of lung cancer, also meeting smoking history criteria with varied compliance, particularly among individuals with a smoking history who are eligible for LDCT.^3^ Screening for other deadly cancers is not recommended for the general population because of their relatively low incidence and harms associated with false positive findings. Risk assessment through a blood test has the potential to increase compliance for cancers for which screening is available and the potential for less common cancers to identify the subset of individuals in the general population who are at sufficiently elevated risk that would benefit from targeted screening.

A multitude of biospecimen types from biological fluids to exhaled breath have been investigated for clinical purposes spanning from risk assessment to early detection of cancer. The spectrum of biomarkers is wide and encompasses products released by tumors or pre-malignant lesions as well as systemic features indicative of host susceptibility or host response to tumor development. ^4^ Biomarkers may be developed for either risk assessment to identify those at increased risk of developing or harboring cancer or for detecting cancer at the time of testing. For common cancers with established screening programs, such as breast or lung cancer, the incidence is sufficiently elevated to justify population-level screening among individuals with adequately elevated risk. For breast cancer, women are considered at elevated risk based on the Gail Model if their 5-year risk is 1.67% or higher. For lung cancer, the estimated risk of those currently eligible under United States Preventive Services Task Force (USPSTF) criteria is 1% over 6 years by the PLCO_m2012_ calculator.^5^ A biomarker test that provides risk assessment for multiple deadly cancers at similar risk thresholds has the potential to identify the smaller subset of individuals in the general population who are at sufficiently elevated risk to warrant targeted screening for the cancer for which they are deemed to be at risk.

A focus of our research is to develop blood-based markers indicative of risk for common and lethal cancers. This is exemplified by our prior findings of a 4-marker protein panel (4MP) consisting of pro-surfactant protein B (ProSFTPB), cancer antigen 125 (CA125), carcinoembryonic antigen (CEA), and cytokeratin-19 fragment (CYFRA) for personalized risk assessment of lethal lung cancer among subjects who have ever-smoked in the Prostate Lung Colorectal and Ovarian (PLCO) Cancer Screening Trial. In this study, the 4MP together with a clinical risk model based on smoking history would have identified for annual screening 9.2% more lung cancer cases while reducing referral by 13.7% among non-cases compared with current USPSTF screening guidelines.^5^ A subsequent study further demonstrated mortality benefit of the 4MP in the PLCO Cohort.^6^ In addition to lung cancer, we have demonstrated that a panel of protein biomarkers informs upon risk of pancreatic cancer up to two years prior to onset of symptoms and clinical diagnosis.^5,7^

Here we report in a blinded validation study, the performance of a multi-cancer risk stratification test (MCaST) that is anchored on the 4MP and that incorporates six additional cancer-associated protein biomarkers for determining risk of developing or harboring nine common and lethal solid cancer types in a prospective lung cancer screening cohort of individuals with a smoking history that were eligible for screening based on current United States Preventive Task Force (USPSFT) criteria.^8^ The cohort consisted of pre-diagnostic plasma samples collected from 73 lung cancer cases and 98 other cancer diagnoses and 3-times more of randomly selected non-case control participants from The Lung Cancer, Early Detection, Assessment of Risk, and Prevention (LEAP) study that did not develop any cancer during the 5-year study period. Assessment of performance included accuracy of cancer signal of origin for those individuals who tested ‘positive’ (i.e. met or exceeded the pre-defined risk thresholds).

## Methods

### LEAP cohort

The Lung Cancer, Early Detection, Assessment of Risk, and Prevention (LEAP) study was an international, prospective, multicenter, longitudinal cohort study launched in 2014 to address needs related to lung cancer screening. Detailed information regarding the LEAP study is reported elsewhere.^8^ Participant information for the LEAP cohort was collected across 10 centers (eight in the United States, one in France, and one in Spain). Participant data, imaging data, and biospecimens were centralized at The University of Texas MD Anderson Cancer Center. Ethical oversight for the study was provided by the Institutional Review Board of The University of Texas MD Anderson Cancer Center (Houston, Texas), which reviewed and approved the study protocols under IRB #2013-0609 and IRB #PA16-0415. The study was conducted in accordance with the Declaration of Helsinki (2013), and all participants provided written informed consent prior to participation.

Briefly, the LEAP study enrolled 2,841 participants who were at elevated risk of developing lung cancer based on the National Comprehensive Cancer Network (NCCN) Guidelines from 8 US centers and 2 international centers (one in France, and one in Spain). The study design included a 2-year active screening phase with annual low-dose chest CT scans, blood biospecimen collection, and optional pulmonary function testing for risk profiling at baseline (T0), year 1, and year 2, with an additional 3-years of follow-up for cancer outcomes.

The current study leveraged pre-diagnostic plasma samples from the 73 lung cancer cases and 98 other cancer cases, as well as 3-times the number of randomly selected non-case control participants (N= 526) from LEAP who did not develop any cancer during the 5-year study period (**Table 1**). For each cancer case and non-case control, all available pre-diagnostic plasma samples (N= up to 3) collected up until the time of a clinical cancer diagnosis or until the end up study follow-up were included (N= 1,235 samples in total). Detailed subject and sample characteristics for lung cancer and other cancer diagnoses in the LEAP cohort stratified by collection sites (US and Europe Sites) are provided in **Supplemental Tables S1-2**.

**Table 1.** Patient and tumor characteristics for the 32 subjects identified to be at elevated risk for common and lethal solid cancer types other than lung cancer based on a positive MCaST in the LEAP cohort.

| **Cancer Type** | **Time from the first positive MCaST to Clinical Diagnosis (Days)** | **Sex** | **Smoking Status** | **Pack Years** | **Stage at time of Clinical Diagnosis** |
| --- | --- | --- | --- | --- | --- |
| Breast Cancer | 528 | Female | Current | 60 | Unknown or unstaged |
| Breast Cancer | 656 | Female | Current | 64.5 | Stage 1 |
| Breast Cancer | 974 | Female | Current | 80 | Unknown or unstaged |
| Breast Cancer | 1770 | Female | Current | 39 | Stage 2 |
| Breast Cancer | 1799 | Female | Current | 49 | Stage 1 |
| Colon and Rectal Cancer | 27 | Male | Current | 35 | Stage 4 |
| Colon and Rectal Cancer | 31 | Male | Current | 48 | Stage 3 |
| Colon and Rectal Cancer | 304 | Male | Current | 32.3 | Stage 2 |
| Colon and Rectal Cancer | 312 | Male | Current | 127.5 | Stage 4 |
| Colon and Rectal Cancer | 317 | Male | Current | 110 | Stage 2 |
| Colon and Rectal Cancer | 1894 | Male | Current | 26.6 | Unknown or unstaged |
| Ovarian Cancer | 697 | Female | Current | 39 | Stage 3 |
| Ovarian Cancer | 903 | Female | Former | 24 | Stage 1 |
| Pancreatic Cancer | 785 | Male | Former | 64.5 | Stage 4 |
| Prostate Cancer | 30 | Male | Former | 72 | Unknown or unstaged |
| Prostate Cancer | 114 | Male | Current | 37.5 | Unknown or unstaged |
| Prostate Cancer | 141 | Male | Current | 39.8 | Unknown or unstaged |
| Prostate Cancer | 372 | Male | Former | 30.8 | Stage 2 |
| Prostate Cancer | 455 | Male | Current | 50 | Stage 2 |
| Prostate Cancer | 462 | Male | Former | 38 | Stage 1 |
| Prostate Cancer | 498 | Male | Current | 43 | Stage 2 |
| Prostate Cancer | 561 | Male | Current | 47 | Stage 2 |
| Prostate Cancer | 677 | Male | Current | 40 | Stage 2 |
| Prostate Cancer | 804 | Male | Current | 39 | Stage 2 |
| Prostate Cancer | 1292 | Male | Current | 25 | Stage 3 |
| Prostate Cancer | 1335 | Male | Former | 49 | Stage 2 |
| Prostate Cancer | 1491 | Male | Current | 55 | Unknown or unstaged |
| Prostate Cancer | 1516 | Male | Current | 64.5 | Stage 3 |
| Prostate Cancer | 1720 | Male | Current | 48 | Stage 1 |
| Prostate Cancer | 8839 | Male | Former | 61.5 | Stage 2 |
| Upper GI Cancer | 224 | Male | Former | 32 | Unknown or unstaged |
| Upper GI Cancer | 600 | Male | Current | 40 | Stage 2 |

### Biomarker assays

The multi-cancer risk stratification test (MCaST) quantifies levels of 10 cancer-associated biomarkers (ProSFTPB, CEA, CA125, CYFRA-21, CA19-9, AFP, HE4, PSA [free and total], and CA15-3) for assessing the risk of developing or harboring any of nine cancer types: lung, prostate, colorectal, breast, ovarian, pancreatic, esophageal, and stomach cancers.

Of the markers, CEA, CA125, CYFRA-21, CA19-9, AFP, HE4, PSA [free and total], CA15-3 were assayed on the Roche Cobas immunoassay platform. ProSFTPB was measured using bead-based immunoassays as previously described^5,6^. Biomarker scores for the lung cancer risk were calculated based on the previously developed logistic regression model ^5,6^. Combination rules and pre-specified clinical decision thresholds for the 9 cancer types are based on a proprietary algorithm established at MD Anderson.

### Statistical analyses

Individualized combination rules based on a broader panel of 10 protein biomarkers for each of the nine specified cancer types were applied using a previously developed proprietary algorithm from MD Anderson using samples independent of LEAP. The MCaST model was calibrated for the general population and cancer-specific risk thresholds for elevated risk were predefined based on established clinical guidelines. For breast cancer, a threshold of ≥0.333% 1-year risk (equivalent to a 1.67% 5-year risk per the Gail model criteria used for identifying women eligible for risk reduction intervention) was used. For lung cancer, a ≥0.167% 1-year risk threshold (equivalent to a 1.0% 6-year risk, consistent with the USPSTF screening eligibility criteria) was applied. The same ≥0.167% 1-year risk cut-off was used for esophageal, gastric, colorectal, pancreatic, liver, ovarian, and prostate cancers, reflecting their similar mortality profile as lung cancer.

Discriminant analyses included time-dependent (e.g. 0-3 months, 0-6 months, 6-12 months, etc) sensitivity and specificity estimates. Performance estimates were calculated at the sample-level and at the subject-level (i.e. a subject is classified as a test ‘positive’ if any of their serial samples show a value exceeding the predefined threshold). Analyses were performed using R software, version 4.5.1 (R Project for Statistical Computing).

## Results

### Predictive performance of the MCaST for lung cancer in the LEAP cohort

In LEAP, 171 cancer diagnoses were made during the 5-year study period. Of these, 118 (69%) were the common and lethal solid cancer types that are the focus of the 10-protein biomarker risk assessment test (*see methods for additional information*). The other 53 (31%) cancer diagnoses were hematological malignancies (N= 13), bladder cancer (N= 13), head and neck cancer (N= 5), kidney cancer (N=4), neuroendocrine tumors (N= 5), and other low-incidence cancer types (N= 13) (*detailed information regarding cancer types are provided in* **Supplemental Tables S1-2**).

Lung cancer comprised 73 (42.7%) of the 171 cancer diagnoses in LEAP, of which 59 (80.8%) were from US sites and 14 (18.9%) were from European sites. Distribution of lung cancer cases by stage at clinical diagnosis were as follows: 42 (57.5%) were stage I; 9 (12.3%) were stage II; 12 (16.4%) were stage III; and 9 (12.3%) were stage IV. For one lung cancer case, staging at time of diagnosis was indeterminant. Median time from blood-collection to a lung cancer diagnosis when considering all sites was 13.52 months (interquartile range [IQR]: 2.31 – 29.6 months).

Among lung cancer cases and when considering sample level data, the overall MCaST positive rate (i.e. ≥ 0.167% 1-year risk threshold for lung cancer risk) was 78.4% (**Figure 1**). The MCaST positive rate increased markedly the closer the blood sample was taken to a clinical diagnosis of lung cancer, with 90% of samples being positive for the 4MP when collected within 3 months of a lung cancer diagnosis, and 85.7% when collected within 1 year preceding a lung cancer diagnosis (**Figure 1**).

**Figure 1.**
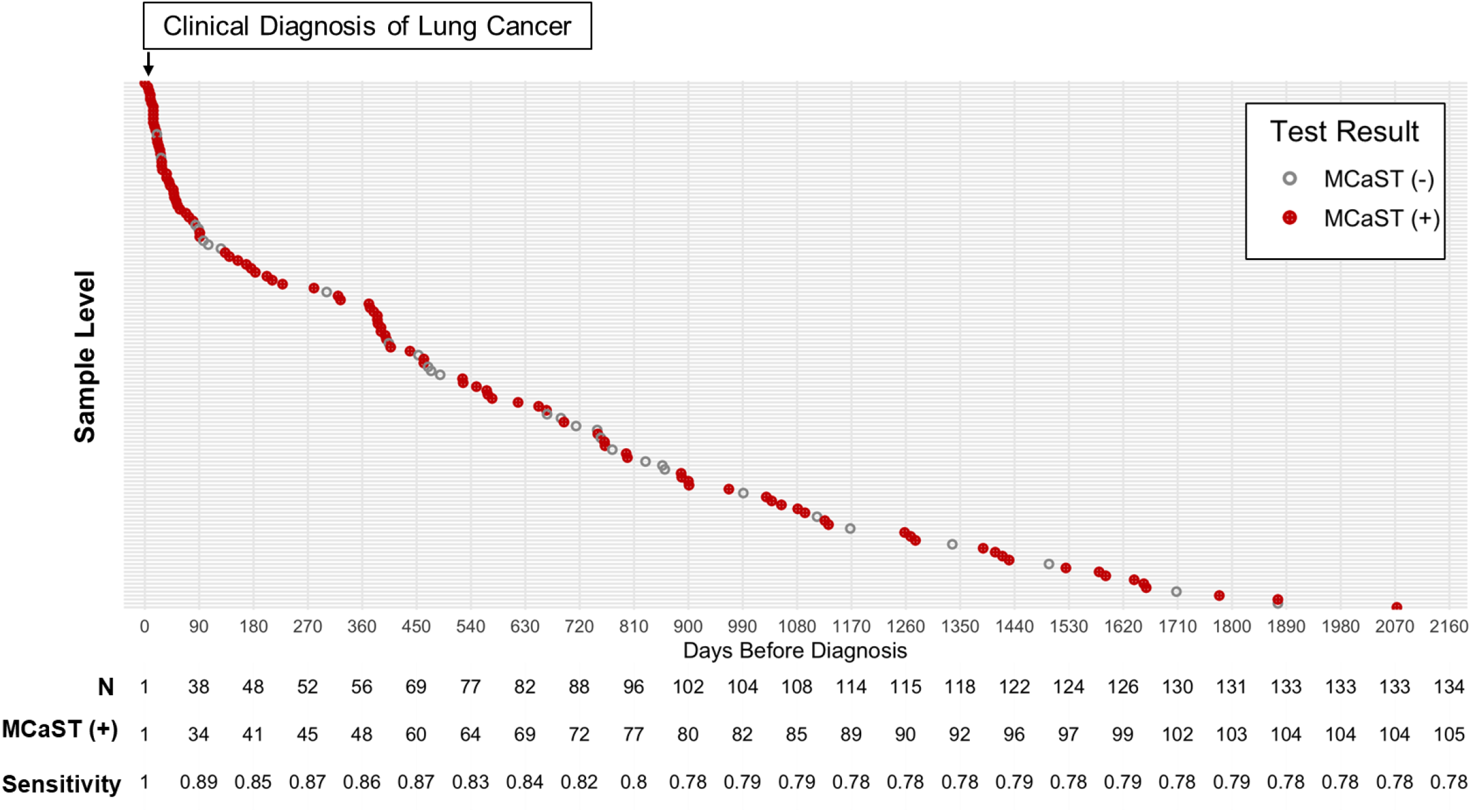
Sample-level MCaST positivity for individuals with a smoking history of ≥20 pack years who went on to receive a lung cancer diagnosis in the LEAP cohort. Y-axis represents sample level information from ever-smokers in the LEAP cohort that went on to receive a lung cancer diagnosis. X-axis depicts time of sample collection to a clinical diagnosis of lung cancer (denoted herein as Time ‘0’). Red nodes represent samples that were positive by the MCaST based on the pre-defined ≥0.187% 1-year risk threshold. Grey nodes represent samples that were MCaST negative. Numbers underneath provides the number of samples collected within each 90-day interval.

At the subject level (i.e. any of serial samples had a MCaST value ≥0.167% 1-year risk threshold), 65 out of the 73 lung cancer cases, including 45 of the 51 early stage (I-II) cases (**Figure 2; Supplemental Figure S1**) were identified as high-risk based on the MCaST. Median (IQR) time from the first positive MCaST to a clinical diagnosis of lung cancer was 12.7 months (1.7 – 27.8 months) (**Figure 2**). Notably, 10 of the 65 subjects that were positive for MCaST had 1-year risk estimates ≥0.833% (equivalent to a 5% 6-year risk), which is consistent with the risk of lung cancer among a symptomatic population (**Supplemental Table S3**).

**Figure 2.**
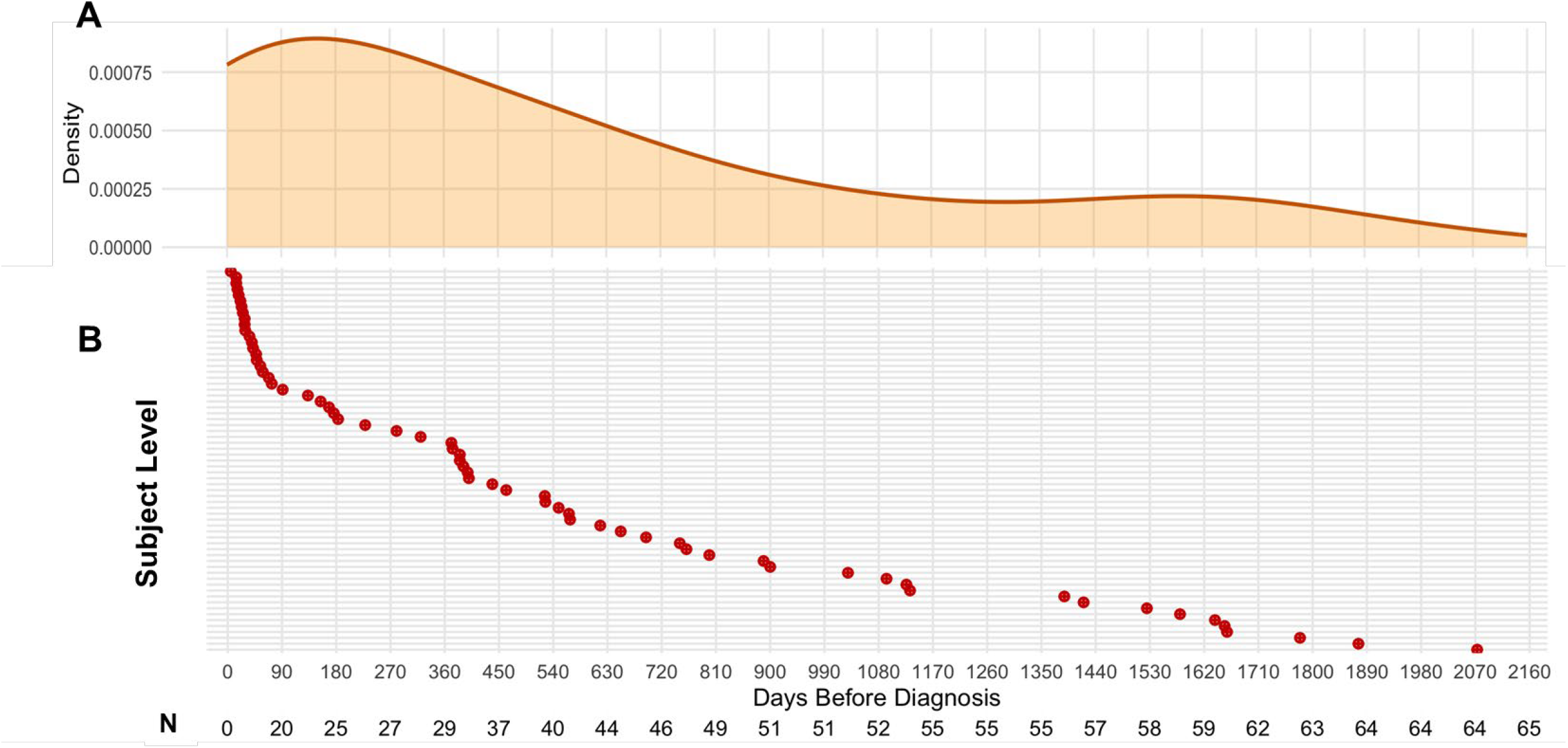
Subject-level MCaST positive rates for individuals with a smoking history of ≥20 pack years who went on to receive a lung cancer diagnosis in the LEAP cohort. **A)** Density plot illustrating the time from the first positive MCaST to clinical diagnosis of lung cancer. **B)** Y-axis reports subject-level information for the 65 lung cancer cases that were identified as high-risk by the MCaST in LEAP based on the pre-defined ≥0.187% 1-year risk threshold. The X-axis depicts the time from the first positive MCaST to a clinical diagnosis of lung cancer (denoted herein as Time ‘0’). Numbers underneath provides the number of samples collected within each 90-day interval.

### Performance of the MCaST for other cancer types

At the sample level, the overall MCaST positive rate for prostate cancer, invasive breast cancer, colorectal cancer, ovarian cancer, pancreatic cancer, and upper GI (esophageal + stomach) cancer based on pre-specified 1-year risk thresholds (0.333% for breast cancer; 0.186% for lung, prostate, colorectal, ovarian, liver, pancreatic, esophageal, and stomach cancers) was 69.6% (**Supplemental Table S3**). When considering samples collected within 1-year, the MCaST positive rate for prostate cancer, invasive breast cancer, colorectal cancer, ovarian cancer, pancreatic cancer, and upper GI cancer cases were 83.3%, 100%, 100%, 50%, 100%, and 66.7%, respectively. For other cancers (excludes lung, prostate, breast, colorectal, ovarian, pancreatic, and upper GI), the sample level MCaST positive rate was 69.3% (**Supplemental Table S3**).

At the subject level, 16 of 18 prostate cancer cases, 5 of 8 invasive breast cancer cases, 6 of 7 colorectal cases, 2 of 4 ovarian cancer cases, 1 of 2 pancreatic cancer cases, and 2 of 3 upper GI (esophagus + stomach) cancer cases (N= 32 cancers in total) were identified as being at elevated risk based on a positive MCaST, corresponding to respective subject-level sensitivity of 88.9%, 62.5%, 85.7%, 50%, 50%, and 67% (**Table 1; Supplemental Table S3 and Figure S2**). Median (IQR) time from the first positive MCaST to a clinical diagnosis of any of the above cancer types was 20.7 months (10.2 – 42.8 months) (**Table 1**). Notably, of the 32 cancer cases that were identified as high-risk by MCaST, 17 (50%) were diagnosed with early-stage (I-II) disease.

### MCaST positive rate among non-case controls

Of the 526 non-case controls, 346 were positive by MCaST (i.e. met or exceed the pre-defined risk thresholds for any one of the 9 cancer types) based on at least one serial samples. Of these, 338 (97.7%) were identified to be at elevated risk for common cancers (lung, colorectal, breast, and prostate cancer) that currently have screening programs, with the majority (N= 288) being at elevated risk of lung cancer.

Of the 346 controls that were positive by MCaST, 160 (46.2%) were at elevated risk for only 1 cancer, 119 (34.4%) were at elevated for 2, and 50 (14.5%) for 3, and 17 (4.9%) were at elevated risk for more than four cancer types (**Supplemental Table S4**).

### Tissue of origin accuracy

At the sample level and when considering cases (any cancer), 250 were MCaST positive (**Supplemental Table S3**). Of these, 100 (40%) were at an elevated risk for only one cancer type, 89 (35.6%) for 2 cancer types, 46 (18.4%) for 3 cancer types, and 15 (6.0%) for more than 3 cancer types (**Figure 3A; Supplemental Table S3**).

**Figure 3.**
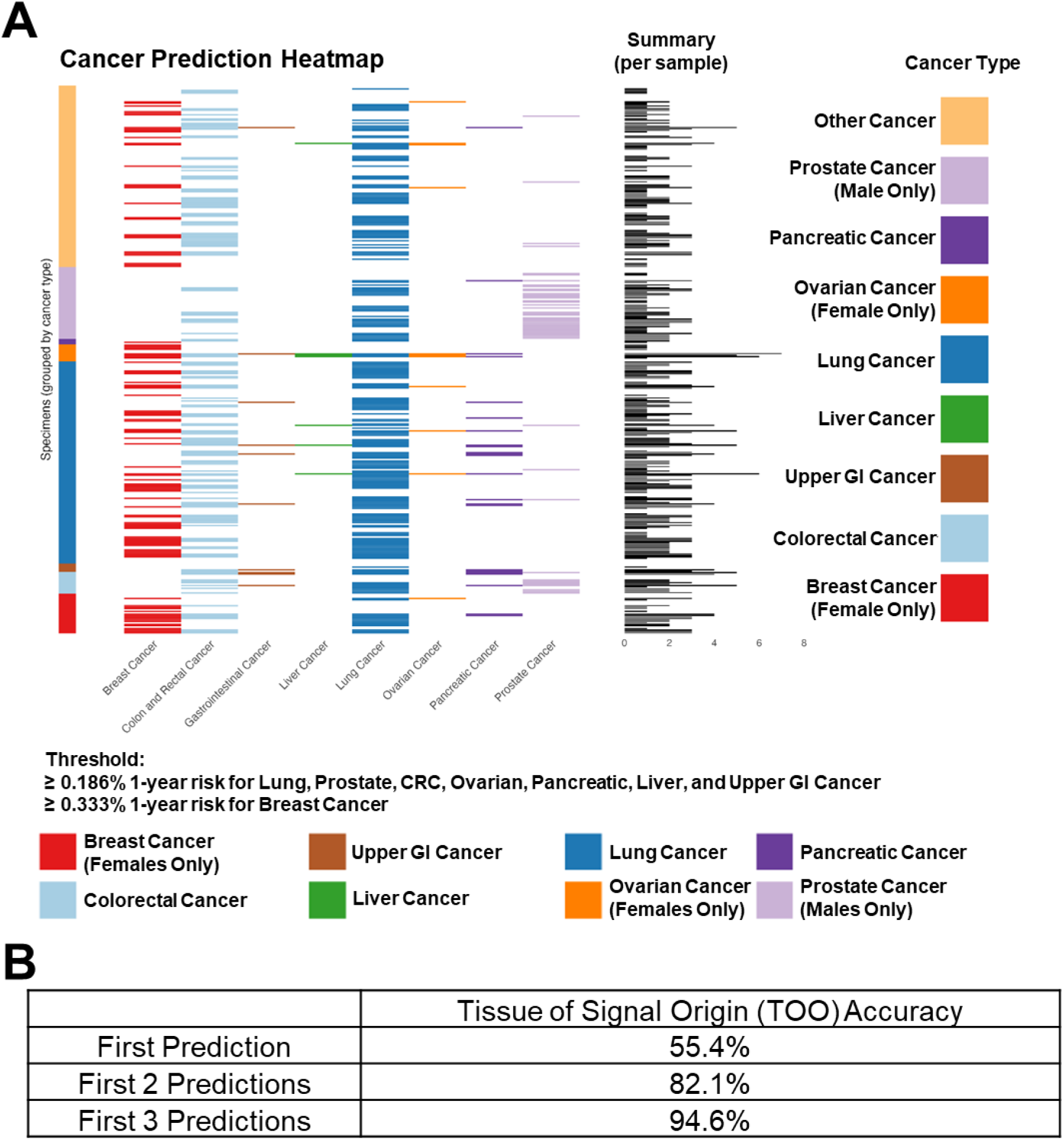
Tissue of signal origin accuracy at the sample level for cases in the LEAP cohort. **A)** Heatmap demonstrating MCaST cancer-specific positive rates (i.e. ≥ 0.187% 1-year risk for lung, prostate, colorectal, ovarian, pancreatic, liver, and upper GI cancer or ≥0.333% 1-year risk for breast cancer) at the sample level (rows) for subjects that went on to receive a cancer diagnosis (cancer-type at diagnosis is defined in colored tiles on the y-axis) in the LEAP cohort. The per sample summary reflects the number of times a given sample was positive (i.e. met or exceed the pre-specified risk threshold) for a given cancer type. **B)** Table representing the tissue of signal origin (TOO) accuracy. For these estimates, the absolute 1-year risk for a given cancer type is considered and ranked from highest to lowest probability. Accuracy is defined by whether a given sample collected from a subject that later went on to receive a cancer diagnosis was accuracy predict based on the first prediction, the first 2 predictions, or the first 3 predictions.

To assess the accuracy of the tissue of origin of the cancer signal (TOO) at the sample level, we evaluated whether the cancer-specific algorithm correctly predicted the cancer diagnosis within the top three predictions, similar to the approach used by others [5, 12], based on ranking of highest of cancer-specific 1-year risk estimates.

The overall TOO accuracy for lung, prostate, breast, colorectal, ovarian, pancreatic, and upper GI (esophageal + stomach) cancers when considering the first predicted cancer type was 55.4%, increasing to 94.6% when considering the top 3 predictions (**Figure 3A-B**). For example, in the context of subjects that went on to develop lung cancer, 55.2% of the samples that tested positive by MCaST had lung cancer as being the highest probability based on 1-year risk estimates, 85.7% had lung cancer within the top 2 highest-risk cancers, and 98.1% had lung cancer within the top 3 highest risk cancers (**Figure 3A**).

## Discussion

We report the performance of MCaST, a protein-based multi-cancer risk assessment test designed to identify individuals at elevated risk for nine common and lethal cancers within a prospective lung cancer screening cohort. The test is anchored on an extensively validated 4MP for lung cancer risk assessment and incorporates six additional cancer-associated proteins to extend its application across prostate, colorectal, breast, ovarian, pancreatic, liver, esophageal, and stomach cancers. Our findings demonstrated that MCaST had high sensitivity for identifying individuals at elevated risk for a broad spectrum of cancers, with 82% of all cancers being identified as high risk, with a median lead time of ∼18 months prior to clinical diagnosis, and a tissue-of-origin accuracy of 94.6% overall with accuracy as high as 98.1% for lung cancer when considering the top three predicted.

A key distinction between MCaST and multi-cancer detection tests, is in intended use. The latter tests are optimized to detect cancers and thus require high specificity for confirmation of presence of cancer at the time of testing.^9-12^ In contrast, MCaST is not designed to detect cancer but rather to identify individuals at elevated risk of developing or harboring a malignancy. This approach inherently prioritizes high sensitivity and risk enrichment, accepting a relatively lower specificity as the intended purpose is to identify individuals who would benefit most from subsequent secondary testing and surveillance.

Whereas the validation study we conducted is limited to individuals with a smoking history that underwent LDCT, it is envisioned that with broader validation, MCaST may have utility for a general population. MCaST would be envisioned as an upstream test in the cancer-screening continuum, to better direct individuals to appropriate clinical work-up. For example, individuals identified to be at elevated risk for lung or colorectal cancer by MCaST would be recommended to undergo guideline-based screening if eligible, whereas those with high-risk profiles for cancers lacking standard screening or ineligible for guideline-based screening may undergo clinically relevant imaging or additional testing with MCD if the tissue of origin is indeterminate with MCaST.

MCaST may also improve adherence to screening programs. Additionally, individuals identified by MCaST at high risk for lung cancer but who do not meet current USPSTF eligibility may be considered for LDCT screening, potentially addressing the limitation that many lung cancers occur in individuals who fall outside of current risk criteria. In prior studies, we demonstrated utility of the 4MP to identify individuals at high risk for lung with a smoking history that do not meet LDCT screening eligibility criteria.^5,6,13,14^ The personalized risk approach has the potential to improve compliance with screening and to identify individuals at risk for cancers for which screening is not available for the general population.

Whereas this study provides strong validation for the MCaST approach, it should be noted that MCaST was positive in approximately 82% of all cancers, but with reduced specificity. Thus, the test is not intended to detect cancer. In the context of the LEAP a proportion of participants were already eligible for standard-of-care screening. However, we note that when excluding individuals eligible for current screening programs, the false-positive rate among remaining participants dropped substantially to approximately 6%, highlighting that the majority of “false positives” represent appropriate identification of subjects already eligible under clinical screening recommendation, with the potential to improve compliance. For cancers other than lung cancer, the number of available pre-diagnostic samples was limited, and larger prospective cohorts will be required to confirm cancer-specific risk models. The LEAP cohort consisted exclusively of individuals with a ≥20PY smoking history that were eligible for lung cancer screening, limiting generalizability to the broader population. Future validation studies are planned among the general population, including never-smokers, to establish the broader utility of MCaST. Further development of MCaST to include additional cancer types may enhance its applicability and value as a broader risk stratification platform. The complementarity of the MCaST with clinical risk prediction models, e.g. PLCO_m2012_ for lung cancer^15,16^ or Gail Model for breast cancer^17^, was not considered. Future studies will evaluate hybrid models that combine biomarker-derived risk with patient characteristics to refine individual risk estimates, potentially improve specificity, reduce unnecessary downstream evaluations, and enhance overall clinical utility across diverse populations.

In conclusion, MCaST represents a biologically grounded tool for personalized cancer risk assessment that we have validated in this study in individuals with a smoking history. Anchored on a validated 4-marker lung cancer panel and extended to encompass nine solid cancers, MCaST demonstrates strong sensitivity, high tissue-of-origin accuracy, and clinical utility as a risk-enrichment test rather than a diagnostic assay for subjects at risk due to their smoking history.

## Data Availability

All data produced in the present study are contained in the manuscript.

## Acknowledgements

We acknowledge the efforts of Dr. Yaxi Li, Ms. Daniela Rodriguez-Perera, Ms. Rachelle Spencer, Ms. Candace Garrett, Ms. Hazel Gudes, Ms. Wenling He, and Ms. Patula Hussein who were involved in sample acquisition and processing as well as assaying of ProSFTPB. The work presented was supported by NIH grant U01CA194733.

## References

1. Caverly TJ, Hayward RA, Reamer E, et al: Presentation of Benefits and Harms in US Cancer Screening and Prevention Guidelines: Systematic Review. J Natl Cancer Inst 108:djv436, 2016

2. Shin WS, Xie F, Chen B, et al: Updated Epidemiology of Gastric Cancer in Asia: Decreased Incidence but Still a Big Challenge. Cancers (Basel) 15, 2023

3. Kim RY, Rendle KA, Mitra N, et al: Adherence to Annual Lung Cancer Screening and Rates of Cancer Diagnosis. JAMA Netw Open 8:e250942, 2025

4. Fitzgerald RC, Antoniou AC, Fruk L, et al: The future of early cancer detection. Nat Med 28:666–677, 2022

5. Fahrmann JF, Marsh T, Irajizad E, et al: Blood-Based Biomarker Panel for Personalized Lung Cancer Risk Assessment. J Clin Oncol 40:876–883, 2022

6. Irajizad E, Fahrmann JF, Marsh T, et al: Mortality Benefit of a Blood-Based Biomarker Panel for Lung Cancer on the Basis of the Prostate, Lung, Colorectal, and Ovarian Cohort. J Clin Oncol 41:4360–4368, 2023

7. Fahrmann JF, Schmidt CM, Mao X, et al: Lead-Time Trajectory of CA19-9 as an Anchor Marker for Pancreatic Cancer Early Detection. Gastroenterology 160:1373-1383.e6, 2021

8. Dennison JB, Gendarme S, Kettner NM, et al: The LEAP Study: A Multicenter Biospecimen and Imaging Resource for Lung Cancer Screening. Cancer Epidemiology, Biomarkers & Prevention, 2025

9. Connal S, Cameron JM, Sala A, et al: Liquid biopsies: the future of cancer early detection. J Transl Med 21:118, 2023

10. Schrag D, Beer TM, McDonnell CH, 3rd, et al: Blood-based tests for multicancer early detection (PATHFINDER): a prospective cohort study. Lancet 402:1251–1260, 2023

11. Lennon AM, Buchanan AH, Rego SP, et al: Outcomes following a false positive multi-cancer early detection (MCED) test: Results from DETECT-A, the first large, prospective, interventional MCED study. Cancer Prev Res (Phila), 2024

12. Lennon AM, Buchanan AH, Kinde I, et al: Feasibility of blood testing combined with PET-CT to screen for cancer and guide intervention. Science 369, 2020

13. Irajizad E, Fahrmann JF, Toumazis I, et al: Biomarker trajectory for earlier detection of lung cancer. EBioMedicine 108:105377, 2024

14. Guida F, Sun N, Bantis LE, et al: Assessment of Lung Cancer Risk on the Basis of a Biomarker Panel of Circulating Proteins. JAMA Oncol 4:e182078, 2018

15. Tammemägi MC, Ruparel M, Tremblay A, et al: USPSTF2013 versus PLCOm2012 lung cancer screening eligibility criteria (International Lung Screening Trial): interim analysis of a prospective cohort study. Lancet Oncol 23:138–148, 2022

16. Choi E, Ding VY, Luo SJ, et al: Risk Model-Based Lung Cancer Screening and Racial and Ethnic Disparities in the US. JAMA Oncol 9:1640–1648, 2023

17. Wang X, Huang Y, Li L, et al: Assessment of performance of the Gail model for predicting breast cancer risk: a systematic review and meta-analysis with trial sequential analysis. Breast Cancer Research 20:18, 2018

