## Supplemental Figures and tables for "Validation of a blood test for multi-cancer risk stratification in a lung cancer screening cohort"

**Table S1.** Subject and specimen characteristics for LEAP US Sites.

| **LEAP US Sites** | **Cases, N** | **Case Samples, N** | **Non-Cases, N** | **Non-Case samples** |
| --- | --- | --- | --- | --- |
| All | 128 | 251 | 336 | 506 |
| Anal Cancer | 1 | 3 | - | - |
| Bladder Cancer | 8 | 20 | - | - |
| Breast Cancer | 6 | 12 | - | - |
| Colon and Rectal Cancer | 4 | 6 | - | - |
| Endocrine Cancer | 2 | 2 | - | - |
| Upper Gastrointestinal Cancer | 3 | 4 | - | - |
| Ovarian Cancer | 3 | 8 | - | - |
| Vaginal Cancer | 1 | 3 |  |  |
| Head and Neck Cancer | 1 | 2 | - | - |
| Kidney Cancer | 3 | 4 | - | - |
| Leukemia | 3 | 7 | - | - |
| Lung Cancer | 59 | 103 | - | - |
| Lymphoma | 8 | 21 | - | - |
| Melanoma | 1 | 3 | - | - |
| Neuroendocrine (NET) tumors | 4 | 9 | - | - |
| Other | 2 | 3 | - | - |
| Pancreatic Cancer | 2 | 4 | - | - |
| Prostate Cancer | 14 | 31 | - | - |
| Throat Cancer | 1 | 2 | - | - |
| Thyroid Cancer | 2 | 4 | - | - |
| Age (IQR) | 65 [62-71] | | 62 [58-67] | |
| Sex | Male: 80, Female: 48 | | Male: 186, Female: 150 | |
| Smoking Status | Current Smoker: 84  Former Smoker: 44 | | Current Smoker: 227  Former Smoker: 109 | |
| Pack Years (IQR) | 46.5 [38.0-63.4] | | 43.0 [35.0-55.0] | |

 IQR: Interquartile Range

**Table S2.** Subject and specimen characteristics for LEAP Europe Sites.

| **LEAP Europe**  **Sites** | **Cases, N** | **Case Samples, N** | **Non-Cases, N** | **Non-Case samples** |
| --- | --- | --- | --- | --- |
| All | 43 | 93 | 190 | 346 |
| Anal Cancer | 1 | 1 | - | - |
| Bladder Cancer | 5 | 11 | - | - |
| Bone Cancer | 1 | 3 | - | - |
| Breast Cancer | 5 | 14 | - | - |
| Colon and Rectal Cancer | 3 | 6 | - | - |
| Ovarian Cancer | 1 | 3 | - | - |
| Head and Neck Cancer | 4 | 7 | - | - |
| Kidney Cancer | 1 | 1 | - | - |
| Lung Cancer | 14 | 31 | - | - |
| Lymphoma | 2 | 3 | - | - |
| Malignant mesothelioma | 1 | 2 | - | - |
| Neuroendocrine (NET) tumors | 1 | 3 | - | - |
| Prostate Cancer | 4 | 8 | - | - |
| Age (IQR) | 61 [56-68] | | 60 [55-65] | |
| Sex | Male: 25, Female: 18 | | Male: 111, Female: 79 | |
| Smoking Status | Current Smoker: 30  Former Smoker: 13 | | Current Smoker: 132  Former Smoker: 58 | |
| Pack Years (IQR) | 40.0 [31.6-50.5] | | 39.0 [31.5-48.0] | |

IQR: Interquartile Range

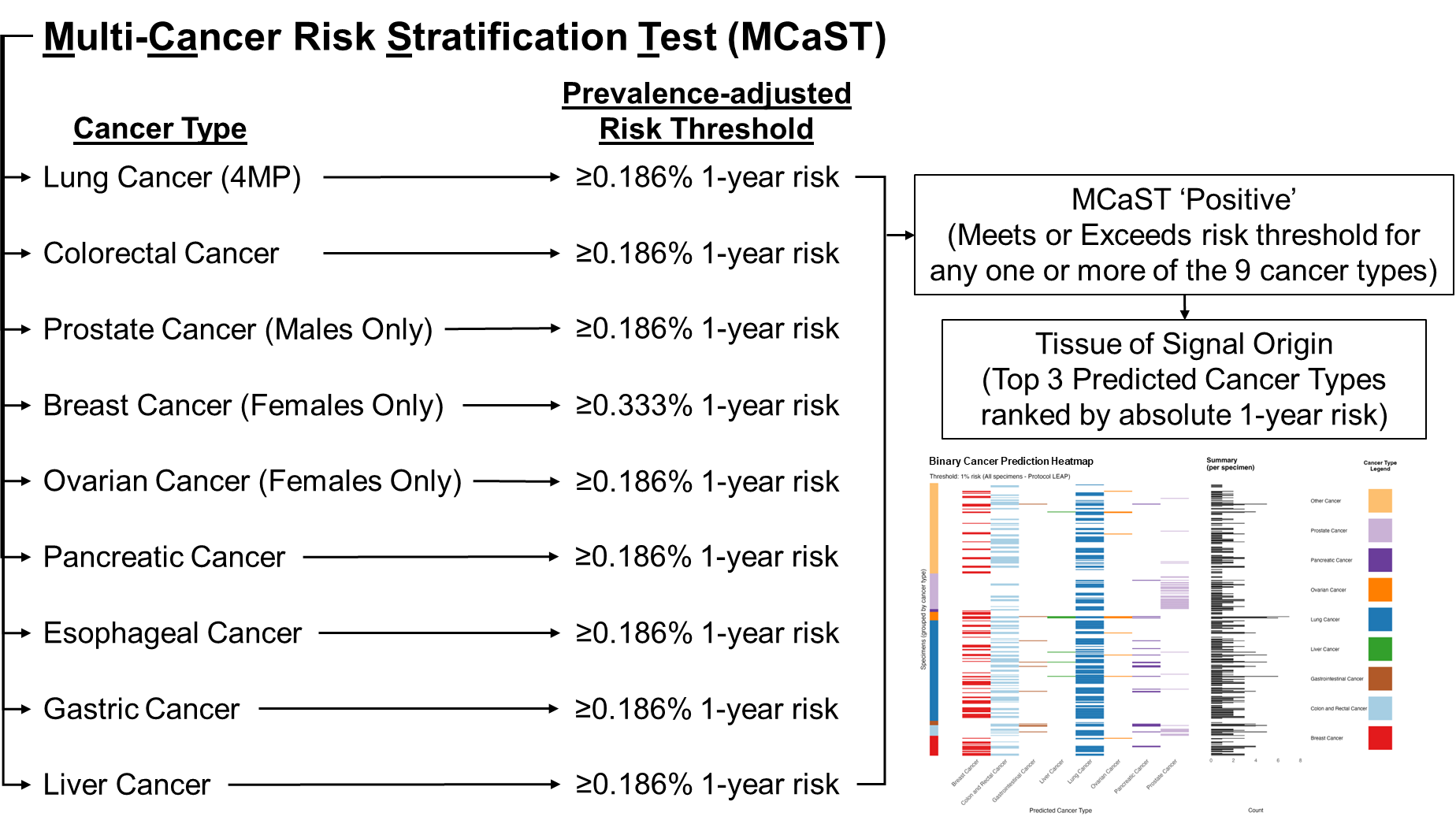

**Supplemental Figure S1. Schematic describing the multi-cancer risk stratification test (MCaST).** The test determines risk of developing or harboring any of nine common and lethal solid cancer types. Individualized combination rules based on a broader panel of 10 protein biomarkers for each of the nine specified cancer types are applied using a previously developed proprietary algorithm from MD Anderson. Cancer-specific risk thresholds are predefined based on established clinical guidelines. A sample is considered ‘positive’ if any one of the 9 cancer types has a value that meets or exceeds the pre-defined risk thresholds. Tissue of Signal Origin (TOO) is defined by rank-order of highest probability based on absolute 1-year risk.
